# Cross-Cultural Adaptation of the Physician Orders for Life-Sustaining Treatment Form in Greece; a pilot cross-sectional descriptive study

**DOI:** 10.1101/2024.08.14.24311981

**Authors:** Dimitrios C. Moustakas, Alexia Bani, Eleni Ntalaouti, Nikolaos Vechlidis, Charalampos Charalampidis, Stamatios Chalvatzis, Sotiria Grigoropoulou, Stylianos Faltsetas, Dimitrios T. Boumpas, Evrydiki Kravvariti, Theodoros Trokanas, Effy Vayena, Sotirios Tsiodras

## Abstract

The current legal framework in Greece does not permit withholding medical interventions to allow natural death, leading to a lack of documented advance directives to guide clinical practice. The aim of this pilot descriptive study is to culturally adapt the Physician Orders for Life-Sustaining Treatment (POLST) form, and use it as a framework with which to assess the readiness of Greek residents for discussions on medical care near the end of life. Patients with severely limited prognosis and/or their family members were interviewed by attending physicians who then recorded their wish with regards to Do-Not-Resuscitate (DNR) status on the adapted POLST form, as well as their level of agreement with signing a similar form. Thirty-one patients with an age range of 18 to 109 years old participated. All had severe prognosis due to various end-stage diseases. Sixteen patients (51.6%) wished for DNR status and didn’t wish for further intervention in case they suffered cardio-respiratory arrest. Presence of an end-stage malignancy was associated with a higher chance of DNR preference (OR: 8.8, 95%CI 1.5-50, p = 0.017) or comfort measures only (OR: 5.6, 95%CI 1.1-30, p = 0.05). This pilot study of patients with severe prognosis hospitalized in a tertiary care center in Greece showed preliminary evidence of high acceptance of natural death and DNR status. POLST forms could be used as a framework for advance care planning through shared decision-making, thus reducing unwarranted treatments and enhancing trust between healthcare professionals and patients at the end of life.

## INTRODUCTION

The application of new technologies in medicine has introduced an array of biomedical interventions, with broad potential to save countless lives (1). Excessive zeal can characterize the medical community’s approach to artificially maintaining a dying patient, employing all available scientific and technological options to achieve a ’victory’ over death, irrespective of the suffering such interventions may cause (2). Herein lies the concept of ‘dysauthanasia’ (dys = difficulty, deprivation // dysauthan = adjective: dying twice; in Latin, dis implies separation and denial). Originally proposed in 1904 by Morcache, dysauthanasia is understood as the preservation of life through disproportionate medical efforts, i.e. as a synonym for therapeutic obsession leading to a prolonged process of death, and physical or psychological suffering (2).

According to data published by the US Centers for Disease Control and Prevention (CDC), the number of inpatient hospital deaths decreased from 776,000 to 715,000 (an 8% decrease), despite an 11% increase in admissions related to advances in palliative and pre-hospital care (3). The most significant decreases in the number of inpatient deaths were observed in patients with end-stage renal failure and cancer patients (4). In a report from Greece 20 years ago, the majority of critically ill patients with a low prognostic index were dying in the hospital setting (5). Specifically, cancer patients in urban centers showed a disproportionately high incidence of in-hospital death, compared to those living in the countryside (6). However, in a small population-based study conducted from 1993 to 2003, only 10% (n=219) of patients with end-stage neoplastic disease expressed a wish to die in hospital (7).

The pandemic caused by the Covid-19 virus brought similar dilemmas concerning end of life support, communication with family members, and the health care provider’s role in decisions about patients in an overburdened healthcare setting – especially in the highest risk elderly patients, who often suffer from multiple comorbidities (8). Due to pandemic public health measures, hospitals were forced to deny visits from relatives, resulting in many patients dying alone, the relatives’ caregiving role abolished (9). Many patients and their relatives elected to refuse intensive care unit (ICU) admission and/or intubation, possibly influenced by prevailing public opinion or misinformation. The concept of making medical decisions based solely on individual needs shifted especially in the intense initial phase of the pandemic, towards an approach that prioritized efficient use of resources, focused on caring for and saving as many patients as possible (10).

End-of life decisions during the pandemic were entangled with serious social challenges introduced by pandemic control measures. Nonetheless, the circumstances brought to the fore the lack of institutional mechanisms for properly recording patient wishes, and the legal vacuum surrounding patient desires for withholding treatment in end-of life situations. This issue, exacerbated during the pandemic, it is a daily reality in hospitals that requires resolution (11). In Greece at present, patient wishes are not formally recorded, with neither inpatient nor outpatient medical records including instructions on life-sustaining medical procedures. Furthermore, the legal framework on allowing natural death is ambiguous, to say the least (12). Recent studies point to the significant impact that respecting patient wishes has on the quality of care, and on compliance of critically ill patients with treatment plans (13).

The aim of this pilot study was to adapt the POLST form for use in the culturally and linguistically diverse context of Greece (14). We sought to evaluate the feasibility and acceptability of implementing the adapted form, assessing factors such as ease of use, time required for completion, perceived utility among healthcare professionals and patients, and related knowledge and attitudes.

## METHODS

### Study design

This pilot cross-sectional descriptive study was performed using the STROBE (Strengthening the Reporting of Observational Studies in Epidemiology (STROBE) Statement guidelines (15). In short, the study involved the cross-cultural adaptation of the copyrighted form of Oregon Portable Orders for Life Sustaining Treatment (POLST) and the pilot study. The adapted POLST form was used to assess attitudes towards allowing natural death, among patients facing severe prognosis in a clinical practice setting at a tertiary care center in Greece.

### Setting and Participants

Eligible participants were all consenting consecutive patients hospitalized at the 4th Internal Medicine Department of the University General Hospital “Attikon” with a severely limited prognosis, based on the Gagné Index for patients over 65 years old, and the Walter Index for patients over 75 years old (16,17). Recruitment was carried out between December 2023 and February 2024.

### Patient involvement

Patients with a severely limited prognosis and their families were not involved in setting the research question or study design. Patients and their families were central to dissemination of the baseline information, which helped to motivate community involvement during and beyond the study.

### Primary outcome

To adapt the POLST form for use in the context of Greece by creating the Attikon POLST Modified Form and to evaluate the feasibility and acceptability of implementing the adapted form in the clinical setting.

### Secondary outcome

To assess factors such as ease of use, time required for completion, perceived utility among healthcare professionals and patients, and related knowledge and attitudes.

### Data management

All data will be anonymised and compliant with regulations including GDPR and ICH Good Clinical Practice (18).

### Sample Size

Sample size was arrived based on similar pilot studies in this field and standard recommendations (19–21).

### Translation and cultural adaptation

The methodology for the POLST form adaptation was based upon the Principles of Good Practice for the Translation and Cultural Adaptation Process for Patient-Reported Outcomes Measures of the International Society of Pharmacoeconomics and Outcomes Research (ISPOR)(22). Each step of the process is described below (See Figure.1)

- Preparation. Decision about which form to use as the source for adaptation. Request and obtain permission to use the copyrighted form of Oregon Portable Orders for Life Sustaining Treatment (POLST) from the Center for Ethics in Health Care at Oregon Health and Science University (OHSU) in Portland, Oregon, United States (23).
- The study was approved by the hospital ethical and scientific board
- Forward Translation. Three native Greek speakers fluent in English performed translations of the source into Greek. Two of those translators were senior medical students and the third was an English philosophy teacher.
- Reconciliation. A team of four healthcare professionals compared and merged the three translations into a single forward translation.
- Back Translation. Two native English speakers fluent in Greek performed independent back translations into the original language.
- Back Translation Review. This version was sent back to OHSU for comparison of the back-translated version of the instrument with the original and revised versions.
- Cognitive Debriefing/Pilot testing and finalization. Form reviewed by eight healthcare professionals who were staff members of the internal medicine clinic and revision of the form followed, according to their comments. Τhen, this pilot cross-sectional study was performed in a sample of 31 severely ill patients with poor prognosis and low expected 1-year survival rate. The questionnaire was completed by the research team at the patient’s bedside in the form of an interview. In the case of a patient’s inability, the interview and data collection was performed with the patient’s next-of-kin or other closed providing usual daily care at home.
- Consultation with an attorney (Athens University professor of law) and presentation of the final form to OHSU.
- Proofreading and final report. Researchers checked the form for minor print errors and produced a detailed report in Greek concerning each adaptation decision made throughout the study.

Our team chose to use the 2023 version of the Oregon POLST form as our primary source, after obtaining permission from the medical director of the Oregon POLST program. While multiple forms are used across the United States, Oregon is the state with the oldest POLST program (24). The investigative team obtained permission on the behalf of the medical director of the Oregon POLST program. The source versions of the Oregon POLST form, the final version of the “Attikon Modified” form adapted to the Greek context, and the back translation are available upon request (Supplementary material Appendix).

The form’s readability level was a concern. To ensure comprehension and accurate understanding, physicians were instructed to administer the completion of the questionnaire at the patient’s bedside in the form of an interview, with the patient or an authorized relative. While in several US states, various healthcare professionals are authorized to sign POLST forms, in Greece advance care planning conversations are considered the responsibility of physicians, and thus only medical doctors were authorized to sign the form. At present, these conversations are not addressed by a specific legal framework, despite the presence of ethics boards in local hospitals of the national health system.

### Statistical analysis

Basic descriptive analysis was performed using SPSS V29 (IBM SPSS Statistics for Windows, Version 29.0, Armonk, NY: IBM Corp.). Patients’ baseline clinical parameters were recorded as absolute numbers and proportions for categorical data, and as median and interquartile ranges for continuous data. Sample size was calculated based on methodology proposed by Rotondi and Donner, as well as previous studies(25–28). All patients approached for study participation accepted and signed a consent form.

## RESULTS

The pilot descriptive study included 31 patients with an age range of 18 to 109 years. A list of medical diagnoses is provided in Table 1. Most participants had underlying end-stage malignancy (32.3%) or a neurological condition (22.6%) and multiple comorbidities (Table 1). According to Gagne and Walter indexes, all included patients had a severe prognosis and high risk of 1-year mortality (16,17). Twenty-seven patients were aged 65 years or older at the time the form was signed, and 64.5 % were female (35.5 % male). Patient residence was categorized as “urban” or “rural”, with a majority (72.2%) coming from urban settings. In most cases, discussion took place with a relative (67,7%).

**Table 1.** Population characteristics and preferences for Cardiopulmonary Resuscitation Orders and Treatment Plan

|  |  |
| --- | --- |
| Person completing questionnaire | N (%) |
| Patient | 6 (19.4) |
| Legally authorized representative | 4 (12.9) |
| Relative, other closed person | 21 (67.7) |
| Underlying condition |  |
| Infection | 5 (16.1) |
| Aspiration | 4 (12.9) |
| Heart failure | 5 (16.1) |
| Malignancy | 10 (32.3) |
| Central nervous system condition (stroke or neurodegenerative disease) | 7 (22.6) |
| Questionnaire data |  |
| Attempt CPR* | 15 (48.4) |
| Do not attempt CPR | 16 (51.6) |
| Comfort measures only | 10 (32.3) |
| Selective treatment | 13 (32.3) |
| Full treatment | 8 (25.8) |

\* CPR: cardiopulmonary resuscitation

Overall, 16 patients (51.6%) wished for DNR status and did not desire further intervention in the case of cardio-respiratory arrest. Of these participants, discussion took place with 12 relatives (57,1%), 1 authorized person, and 3 patients. Fifteen patients (48.4%) wished to attempt resuscitation (Table 1). Furthermore, 13 patients (41.9%) wished for Selective Treatment Orders (generally avoid the intensive care unit), 10 (32.3%) for Comfort Measures Only (patient prefer no transfer to hospital for life-sustaining treatments. Transfer if comfort needs cannot be met in current location), and 8 (25.8%) for Full Treatment Orders (transfer to hospital and/or intensive care unit, if indicated) (Table 1). Over the course of the interviews, none of the four interviewers reported a lack of clarity in completing the form. Statistical significance for patients desiring or not desiring intervention is shown in Table 2. Presence of an end-stage malignancy was associated with a higher preference for DNR (OR: 8.8, 95%CI 1.5-50, p = 0.017) or comfort measures only (OR: 5.6, 95%CI 1.1-30, p = 0.05). Statistical analysis of differences between urban and rural status, age, and comorbidity status did not reveal noteworthy differences.

**Table 2.**
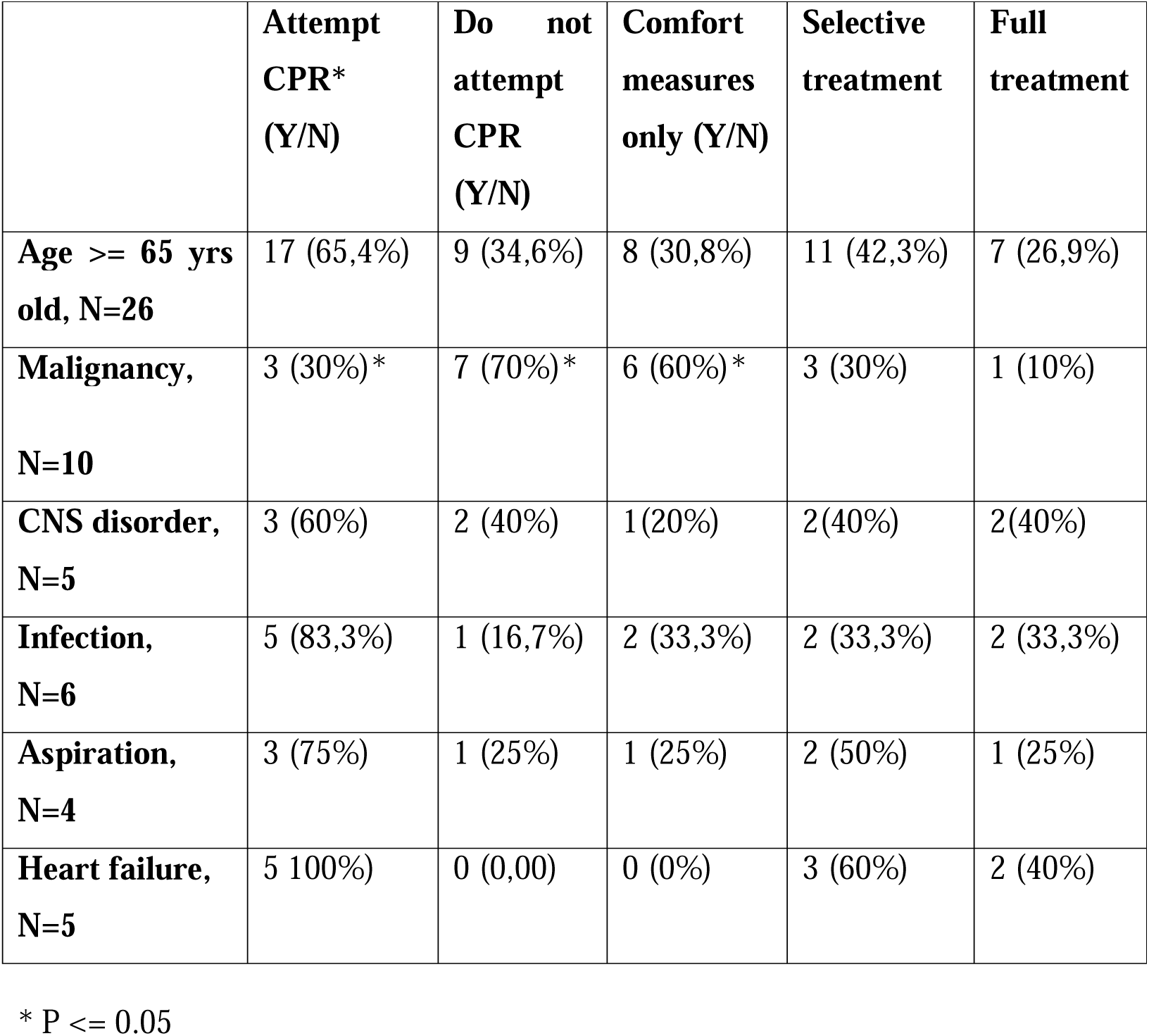
Covariates examined regarding end-of-life decision. Statistical significance for patients desiring or not desiring intervention is shown with an asterisk. For continuous variables we used independent samples t – test, for categorical chi square and risk estimate

|  | **Attempt**<br>**CPR\***<br>**(Y/N)** | **Do not**<br>**attempt**<br>**CPR**<br>**(Y/N)** | **Comfort**<br>**measures**<br>**only (Y/N)** | **Selective**<br>**treatment** | **Full**<br>**treatment** |
| --- | --- | --- | --- | --- | --- |
| **Age >= 65 yrs**<br>**old, N=26** | 17 (65,4%) | 9 (34,6%) | 8 (30,8%) | 11 (42,3%) | 7 (26,9%) |
| **Malignancy,**<br>**N=10** | 3 (30%)* | 7 (70%)* | 6 (60%)* | 3 (30%) | 1 (10%) |
| **CNS disorder,**<br>**N=5** | 3 (60%) | 2 (40%) | 1(20%) | 2(40%) | 2(40%) |
| **Infection,**<br>**N=6** | 5 (83,3%) | 1 (16,7%) | 2 (33,3%) | 2 (33,3%) | 2 (33,3%) |
| **Aspiration,**<br>**N=4** | 3 (75%) | 1 (25%) | 1 (25%) | 2 (50%) | 1 (25%) |
| **Heart failure,**<br>**N=5** | 5 100%) | 0 (0,00) | 0 (0%) | 3 (60%) | 2 (40%) |

\* P <= 0.05

## DISCUSSION

To our knowledge, this is the first study to explore the possible implementation of Portable Orders for Life–Sustaining Treatment in Greece. Αs a component of advance care planning, POLST could increase engagement in the therapeutic plan as well as increase trust between patients and doctors (29,30). The 2014, the report ‘Dying in America’, by the Committee on Approaching Death: Addressing Key End of Life Issues; Institute of Medicine, suggested that all states have the responsibility to create a POLST program (31). POLST is a method of recording and honoring patient’s attitudes regarding DNR orders and selective treatment options (32,33). According to Sinclair C., “the phrase ‘do not resuscitate’ signals an intent to withhold or refuse. It says you’re not going to do something. To ‘allow natural death,’ on the other hand, connotes permission. It doesn’t sound so overwhelming or scary” (34). A non-randomized study from Hahnemann University Hospital in Philadelphia, reported that 78-83% of healthcare professionals would consent to an “Allow Natural Death” (AND) order if a requested patient was near death (35).

In many countries, DNR orders are not legally binding, and thus, fulfillment of patient wishes is not guaranteed. Studies from several European countries have found that the preference for withdrawal of therapy is more common in northern Europe (Denmark, Finland, Ireland, Sweden, Switzerland, England, Scotland, Belgium and the Netherlands) than in southern Europe (Greece, Italy, Portugal, Israel, Spain and Turkey) (36–38).

The development of legal treatment of POLST in Greece would involve the Convention of the Council of Europe for the Protection of Human Rights and Dignity of the Human Being, with regard to the Application of Biology and Medicine: Convention on Human Rights and Biomedicine (hereinafter the Oviedo Convention)(39), the Greek Code of Medical Deontology (hereinafter CMD), and the Greek Penal Code.

Article 9 of the Oviedo Convention, ratified with Greek Law 2619/1998, provides that “The previously expressed wishes relating to a medical intervention by a patient who is not, at the time of the intervention, in a state to express his or her wishes shall be taken into account”. This provision appears to be limited to living wills, thus excluding health-care proxy directives from its scope. Furthermore, the wording “shall be taken into account” is legally ambiguous concerning the binding effect in signatory member states. The issue is further clouded by §62 of the Explanatory Report to the Oviedo Convention, which acknowledges that “taking previously expressed wishes into account does not mean that they should necessarily be followed”. In this way, and without explicit mention in the legal text, the possibility of disregarding a patient’s wishes is introduced. The Oviedo Convention seemingly does not directly affect the legal force of living wills at a national level, especially in signatories where living wills are not legally binding (e.g. Greece), while it accepts medical paternalism to a certain extent in the area of advance directives.

On the other hand, DNR orders constitute an issue of penal law within the ambit of the Greek Penal Code; in particular its Articles 299, 300 and 301, which criminalize intentional homicide, ‘consensual homicide’ (euthanasia), and ‘suicidal complicity’, respectively (in the case of a DNR order by omission)(40).

Greek Law 3418/2005, which integrated the Code of Medical Deontology, regulates solely palliative care as an end-of-life choice for terminally ill patients. Specifically, according to Article 29 § 1 CMD “At the final stage of an incurable illness, even if all treatment margins have been exhausted, a doctor shall ensure the psychosomatic pain relief of the patient. The doctor offers palliative care and collaborates with his or her close members in this direction. In any case, the doctor shows compassion to the patient until the end of his life and ensures that the patient maintains his dignity until this point”. Introducing the notion of ‘futile treatment’ (or ‘therapeutic obstinacy’), this article accepts the occurrence of death, without hastening it. However, the article does not treat the patient by ‘allowing to die’ or ‘letting die’, which entails an intentional avoidance of causal intervention (e.g. CPR), so that disease, system failure or injury causes death. For this reason, Article 29 § 1 CMD does not clash with Articles 299, 300 and 301 of the Greek Penal Code, which criminalize acts of ‘killing’ (40).

The adaptation of the POLST form for the Greek context, along with the creation of the “Attikon Modified” POLST form, require compliance with the Greek law and regulations, and staff education concerning those laws. The methodological steps taken were in accordance with ISPOR; the guidance of principles for future adaptation of POLST to other countries by Mayoral F.S. et al. 2018; and the Consensus-based Standards for the Selection of Health Status Measurement Instruments (COSMIN) checklist.(17,23,37–39).

Our pilot study consisted of 31 patients with a poor prognosis, primarily aged 65 and over. Despite this small sample, we can offer some useful preliminary observations. Ιn Greek hospitals, decisions at the end of life are commonly discussed with relatives (67,7%). If DNR alternative was offered, 51,6% of the patients and family members commented that they would choose this option. More than half of the relatives 57,1% expressed preference for a for DNR status. If this preference were to hold true in a larger study, it would present a significant point of concern for the healthcare system, given that at present, a DNR option is not available to patients in Greece. Notably, full life-saving treatment was in accordance with the wishes of only 25,8% of the patients and relatives.

The present study has several limitations. First, none of the translators were certified professional translators. The existing 1-year mortality risk scores show moderate prognostic performance, discriminatory power and calibration, when used in older adults with multiple comorbidities (40). The study was restricted to one hospital in Athens. However, cross-cultural adaptation could be considered equal for a country in which one language is spoken across all regions. The POLST form provides a foundation for future research and implementation efforts not only within Greece, but also for other Greek-speaking regions such as Cyprus.

## CONCLUSION

Our pilot study, sheds light on critical aspects of end-of-life decision-making in modern Greek healthcare settings. The absence of a legally binding DNR option prevents medical professionals from fulfilling patient preferences and promoting the best quality of care at the end of life. Despite the study’s limitations, including its single-site focus, the findings offer valuable insights into the current landscape of end-of-life care in Greece. Moving forward, continued efforts are needed to enhance staff training, expand research scope, and engage stakeholders in advancing end-of-life care practices. Improvement of the current legal framework through establishment of a new framework is of tremendous importance.. By addressing these challenges and building upon our findings, we can strive towards a more patient-centered approach to end-of-life care that respects individual preferences, fosters trust between patients and healthcare providers, and upholds the principles of dignity and autonomy in healthcare decision-making.

## Data availability statement

The authors confirm that the data supporting the findings of this study are available within the article and its supplementary materials.

## Data Availability

All data produced in the present study are available upon reasonable request to the authors

## Acknowledgements

The authors would like to thank Dr. Mullowney, Medical Director of the Oregon POLST Program, Chair of the Oregon POLST Coalition and Assistant Professor of Medicine, Division of General Internal Medicine and Geriatrics, School of Medicine at OHSU (Oregon Health & Science University) for her helpful advice and comments. We would also like to thank Shannon Hubbs for her editorial assistance in the preparation of the manuscript.

Appendix

---

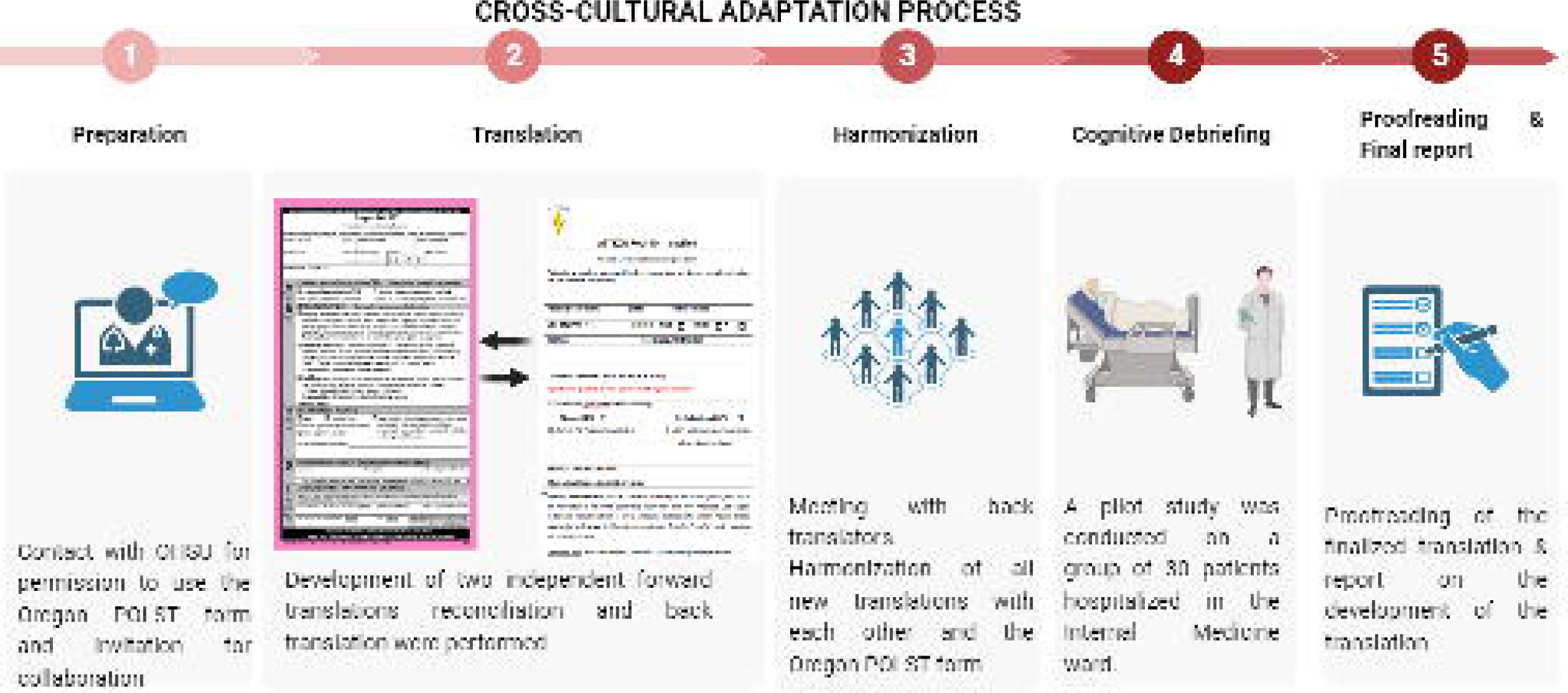

## Notes

### Competing Interest Statement

The authors have declared no competing interest.

### Funding Statement

This study did not receive any funding

### Author Declarations

The study was approved by the University General Hospital Attikon ethical and scientific board, Rimini 1, Haidari, Athens, Greece

